# Unraveling the Decline: Search for Factors Behind the Trend in Parkinson’s Disease Incidence Rates – A Cohort Study of German Health Claims Data

**DOI:** 10.1101/2023.06.29.23291993

**Authors:** Fink Anne, Maria Angeliki S. Pavlou, Roomp Kirsten, Doblhammer Gabriele, Jochen G. Schneider

**Affiliations:** German Center for Neurodegenerative Diseases (DZNE), Bonn, Germany; Department of Life Sciences and Medicine (DLSM), University of Luxembourg, 6, avenue du Swing, L-4367 Belvaux, Luxembourg; Luxembourg Centre for Systems Biomedicine and Department of Life Sciences and Medicine, University of Luxembourg, University of Luxembourg, Esch-sur-Alzette, Luxembourg; Institute for Sociology and Demography, University of Rostock, Rostock, Germany; Saarland University Medical Center, Departments of Internal Medicine II and Psychiatry, Homburg Saar, Germany

## Abstract

**Importance:** Sporadic Parkinson’s disease (PD) is the second most common neurodegenerative disease in older age. Recent studies have led to a debate regarding trends in incidence and prevalence.

**Objective:** We aimed to investigate age-specific incidence rates of PD and possible explanations for the observed trend in Germany.

**Design:** We studied two randomly selected longitudinal cohorts, each consisting of 250,000 individuals aged 50 and above.

**Setting:** We started observing them at the beginning of 2004 and 2014, and followed them through the end of 2009 and 2019, respectively. We compared age-specific incidence rates for both cohorts and performed Cox regression models to calculate the hazard ratios (HR) of PD in the second period compared with the first period ten years earlier, adjusted for age, sex, and several prodromal factors, comorbidities, and risk factors of PD.

**Main outcome and Measures:** Main outcome was the age-specific incidence trends of Parkinson’s disease (PD) in two cohorts using routine health claims data

**Results:** For each age-group in men and women, we found lower age-specific PD incidence rates in the second period, except for the lowest age-group (50-54) in men. Cox regression analysis demonstrated an overall 20% risk reduction of PD incidence (HR=0.80, 95% confidence interval [0.75-0.86]). Mean age at diagnosis increased in men (+1.98 years) and women (+0,87 years). Stepwise adjustment for prodromal symptoms, comorbidities, and risk factors revealed a 22%-49% risk reduction of PD incidence. Sensitivity analysis considering the competing event of death showed a 48% risk reduction (HR=0.52 [0.48-0.56]), demonstrating the independence of the time trends from changes in death rates.

**Conclusion:** Our data show that the risk of PD has decreased over time, and that this decrease is independent of factors such as changes in death rates, age structure, sex, motor and sensory impairments, sleep and psychiatric conditions, comorbidities, and specific risk factors.

**Key points:** *Question:* While several studies have reported on the incidence and prevalence of PD, recent data from Germany have sparked a debate regarding the validity and interpretation of these trends. This study aims to investigate the age-specific incidence rates of Parkinson’s Disease (PD) and explore possible explanations for the observed reduced trend in Germany.

*Findings:* We assessed data from a longitudinal cohort from 2004 to 2009 and 2014 to 2019, respectively. By comparing age-specific incidence rates between the two cohorts we reveal a consistent reduction in the age-specific incidence rates of PD in the second period compared to the first period, except for the lowest age-group (50-54)

*Meaning:* While several studies have reported on the incidence and prevalence of PD, we identified intriguing trends that warrant further investigation.

## Introduction

Parkinson’s disease (PD) is a progressive degenerative disorder of the central nervous system, predominantly affecting older adults and with multifactorial origins. Early symptoms of PD, such as constipation, sleep abnormalities, anemia, parosmia, skin disorders, and diabetes, can occur decades before the onset of the disease ^1^. Overt disease is characterized by tremors, muscle rigidity, and slowness of movement ^2^. In 2019, it was reported that approximately 8.5 million people may live with PD worldwide. The prevalence of the disease has doubled in the last 25 years, with further increase projected until the year 2030 ^3,4^.

In Germany, a country with a rapidly aging society, PD disease burden is expected to increase substantially. In fact, one national study which examined insurance claims data from 2011 to 2019, found an increase in the absolute number and increased morbidity and mortality in PD patients as compared to persons without PD in a predominantly rural area of Germany ^5^. Recent cross-sectional study data from 2010 to 2019 reported a stable prevalence of PD patients and a slight decrease in new PD patients relative to all patients in general practitioners and specialist offices, indicating no relative increase alongside the absolute numbers ^6^. These data were complemented by a recent nationwide assessment of data obtained from pseudonymized, cross-insurance claims data examining the years 2013 to 2019 yielding successive decreases in annual incidence rates between 25-30%. This decrease was observed in all age groups above 50 years, independent of sex, and was accompanied by an overall lower proportion of women suffering from PD over time ^7^. The latter report initiated a debate among specialists regarding the validity of the data^8^.

Prompted by the inconclusive results and ensuing debate, we assessed age-specific incidence trends of Parkinson’s disease (PD) in two cohorts using routine health claims data from Allgemeine Ortskrankenkasse (AOK), Germany’s largest health insurer. We examined how changes in age and sex structure, prodromal and risk factor composition, and mortality could explain the observed trends.

## Material and Methods

### Data

We used two random samples of members of Germany’s largest public health insurance fund, the Allgemeine Ortskrankenkassen (AOK), each consisting of 250,000 people aged 50 years and over at the beginning of 2004 and 2014. The claims data were anonymized and we did not have access to the primary data, hence no ethical review or patient consent was required. The individuals were followed until the end of 2009 and 2019, respectively. In addition to demographic information on sex, year and month of birth/death, and region of residence, our data include inpatient and outpatient medical diagnoses coded according to the 10th revision of the International Statistical Classification of Diseases and Related Health Problems (ICD-10): https://apps.who.int/iris/handle/10665/246208.

To create our analysis sample, we identified new PD cases in both cohorts, excluding individuals with valid PD diagnoses in 2004/05 and 2014/15. The first PD diagnosis between 2006-2008 or 2016-2018 was considered, while 2009 and 2019 were used for diagnosis validation. PD was defined using ICD-10 code G20. Our internal validation strategy involved inpatient discharge or secondary diagnoses and verified outpatient diagnoses. Valid diagnoses required concurrent inpatient and outpatient diagnoses or a second occurrence over time. In the case of death in the quarter of the first diagnosis, it was considered valid even without a confirmed diagnosis.

### Statistical analysis

To compare the incidence rates of PD between the two periods, we calculated age-specific PD incidence rates by dividing the number of PD cases per 1,000 person-years at risk by sex and period. We smoothed the frequency distribution of age at first PD diagnosis using the Stata smoother “4253eh,twice” and performed Cox proportional hazard regression models and calculated the PD hazard ratios (HRs). At first, we performed a regression model with period as an explanatory variable to quantify the time trend effect (model 1), adjusted only for sex and age, modelled by a second-degree polynomial function (age+age^2^). Secondly, we ran several regression models additionally adjusted for motor, sensory and autonomic presentations associated with PD (tremor G25.0, G25.1, G25.2, R25.1; gait impairment R26; anosmia R43.0; change in skin sensation R20.0, R20.1, R20.2, R20.3; constipation K59.0; dizziness R42; model 2), sleep and psychiatric presentations (restless legs syndrome G25.80, G25.81; sleep apnea G47.3; parasomnia F51.3, F51.4, F51.5, G47.8; depression F32, F33; schizophrenia F20; bipolar disorder F31; model 3), typical comorbidities of PD (epilepsy G40; migraine G43; gastroesophageal reflux disease K21; gastritis K29; Crohn’s disease K50; seborrheic dermatitis L21; model 4), or risk factors associated with PD (type 1 diabetes E10; type 2 diabetes E11; hypertension I10; hypercholesterolemia E78; traumatic brain injury S06.0, S06.1, S06.2, S06.3; alcohol misuse F10.1, F10.2; model 5). The full model (model 6) included all prodromal features, comorbidities, and risk factors mentioned above. The selection of covariates was based on a recent study showing a significantly higher prevalence in PD patients than in healthy controls several years prior to the first PD diagnosis ^2^. The analysis started on 1st January 2006/2016 and was measured in months.

The time of PD diagnosis was set in the middle of each quarter. The time of death was defined as the middle of the month of death. Individuals were censored at the time of change in public health insurance or at the end of the follow-up period (end of 2008/2018). The proportionality assumption was tested for the Cox models.

For the sensitivity analysis, we performed Fine and Gray competing risk regression analysis of the full model ^9^, considering death as a competing event (model 7).

All analyses were performed using Stata/MP 16.1.

## Results

### Descriptive results

Our analysis sample consisted of 451,467 persons free of PD at the beginning of the observation period, in 2006 (n=224,366) and 2016 (n=227,101). The sex and age distributions of both cohorts are presented in Table 1. Of these, 1,786 individuals developed PD between 2006 and 2008 and 1,514 individuals developed PD between 2016 and 2018. Apart from the lowest age group (50-54 years) in men, every age-specific PD incidence rate in the second period was lower than that 10 years earlier. Significant decreases were observed in the 60-64 and 70-74 age groups in men and in the 80-84 age group in women. As expected, the incidence of PD increased with age up to the age group 85 to 89 years. The highest age group (> 90 years) had a slightly lower incidence rate. A detailed table with the incidence rates is provided in the Supplementary Appendix (see Table S1).

**Table 1:** Baseline demographic characteristics of the two Parkinson’s disease (PD)-free cohorts. Source AOK 2004-2009, AOK 2014-2019.

| Demographics | 2006 |  | 2016 |  | Total |  |
| --- | --- | --- | --- | --- | --- | --- |
|  | No. | (%) | No. | (%) | No. | (%) |
| Gender |  |  |  |  |  |  |
| Men | 94,971 | (42.3) | 101,133 | (44.5) | 196,104 | (43.4) |
| Women | 129,395 | (57.7) | 125,966 | (55.5) | 255,361 | (56.6) |
| Age group |  |  |  |  |  |  |
| 50-54 | 26,020 | (11.6) | 25,505 | (11.2) | 51,525 | (11.4) |
| 55-59 | 30,563 | (13.6) | 39,291 | (17.3) | 69,854 | (15.5) |
| 60-64 | 29,550 | (13.2) | 33,359 | (14.7) | 62,909 | (13.9) |
| 65-69 | 40,982 | (18.3) | 28,403 | (12.5) | 69,385 | (15.4) |
| 70-74 | 34,180 | (15.2) | 25,469 | (11.2) | 59,649 | (13.2) |
| 75-79 | 28,101 | (12.5) | 32,193 | (14.2) | 60,294 | (13.4) |
| 80-84 | 19,855 | (8.9) | 22,423 | (9.9) | 42,278 | (9.4) |
| 85-89 | 8,994 | (4.0) | 13,689 | (6.0) | 22,683 | (5.0) |
| 90+ | 6,121 | (2.7) | 6,769 | (3.0) | 12,890 | (2.9) |
| Total | 224,366 | (100.0) | 227,101 | (100.0) | 451,467 | (100.0) |

Over the 10 years, the mean age at first PD diagnosis increased significantly from 74.57 years [95% CI:74.01-75.13] to 76.55 years [75.95-77.15] in men and from 77.94 years [77.43-78.45] to 78.81 years [78.20-79.41] in women. Figure 2 shows the observed and smoothed distribution of age at the first PD diagnosis in men and women in both periods. For both sexes, there was a marked decrease in new cases of PD between the ages of 65 and 75. This is not only due to a possible lower disease burden but also to a cohort effect, as these age groups of the second cohort come from smaller birth cohorts (see also Table 1 for the number of individuals in each age group). For men, the curve in the second period shows a significant shift to higher ages, whereas for women, the distribution is narrower in the second period.

**Figure 1:**
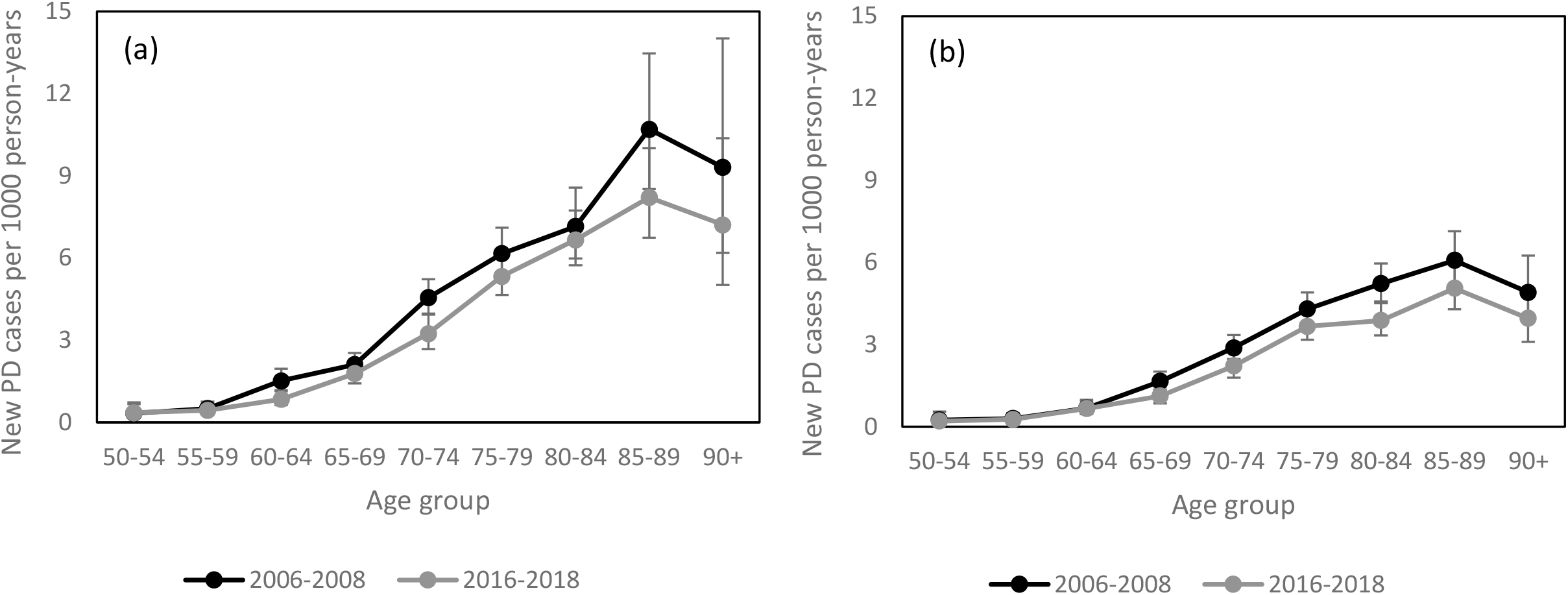
Age-specific Parkinson’s disease (PD) incidence rates per 1,000 person-years for the two periods 2006-2008 and 2016-2018, in men (a) and women (b). Source AOK 2004-2009, AOK 2014-2019.

**Figure 2:** Observed and smoothed frequency distribution of age at first diagnosis of Parkinson’s disease (PD) for the two periods 2006-2008 and 2016-2018, in men (a) and women (b). Source: AOK 2004-2009, AOK 2014-2019.

### Model results

Controlling for age and sex, Cox regression revealed a 20% reduced risk of PD (HR=0.80 [0.75-0.86]) in the 2016-2018 period as compared to the first period ten years earlier (Table 2, model 1). Men had a significantly increased risk of PD (HR=1.55 [1.44-1.66]) compared to women. The estimators for age showed a positive association with the linear term (HR=1.02 [1.01-1.03]) and a negative association with the quadratic term (HR=0.997 [0.997-0.997]), representing the leveling-off of PD incidence in the highest ages.

**Table 2:** Cox regression models for the risk of Parkinson’s disease (PD) in terms of hazard ratios (HR). Source AOK 2004-2009, AOK 2014-2019.

| Variable | Model 1 |  |  | Model 2 |  |  | Model 3 |  |  | Model 4 |  |  | Model 5 |  |  | Model 6 |  |  |
| --- | --- | --- | --- | --- | --- | --- | --- | --- | --- | --- | --- | --- | --- | --- | --- | --- | --- | --- |
|  | HR | P value | 95% CI | HR | P value | 95% CI | HR | P value | 95% CI | HR | P value | 95% CI | HR | P value | 95% CI | HR | P value | 95% CI |
| **Period (Ref.: 2006-2008)** |  |  |  |  |  |  |  |  |  |  |  |  |  |  |  |  |  |  |
| 2016-2018 | 0.80 | <0.001 | 0.75 0.86 | 0.57 | <0.001 | 0.53 0.61 | 0.67 | <0.001 | 0.62 0.72 | 0.78 | <0.001 | 0.73 0.84 | 0.77 | <0.001 | 0.72 0.83 | 0.51 | <0.001 | 0.47 0.55 |
| **Sex (Ref.: Women)** |  |  |  |  |  |  |  |  |  |  |  |  |  |  |  |  |  |  |
| Men | 1.55 | <0.001 | 1.44 1.66 | 1.67 | <0.001 | 1.55 1.79 | 1.89 | <0.001 | 1.76 2.03 | 1.54 | <0.001 | 1.43 1.65 | 1.53 | <0.001 | 1.43 1.64 | 1.91 | <0.001 | 1.77 2.05 |
| Age | 1.02 | <0.001 | 1.01 1.03 | 1.00 | 0.630 | 0.99 1.01 | 1.02 | <0.001 | 1.01 1.02 | 1.02 | <0.001 | 1.01 1.03 | 1.02 | <0.001 | 1.01 1.03 | 1.00 | 0.776 | 0.99 1.01 |
| Age<sup>2</sup> | 0.997 | <0.001 | 0.997 0.997 | 0.997 | <0.001 | 0.997 0.997 | 0.997 | <0.001 | 0.997 0.997 | 0.997 | <0.001 | 0.997 0.997 | 0.997 | <0.001 | 0.997 0.998 | 0.997 | <0.001 | 0.997 0.998 |
| **Motor, sensory, and autonomic presentations** |  |  |  |  |  |  |  |  |  |  |  |  |  |  |  |  |  |  |
| Tremor | NA |  |  | 11.31 | <0.001 | 10.36 12.35 | NA |  |  | NA |  |  | NA |  |  | 9.70 | <0.001 | 8.88 10.61 |
| Gait impairment | NA |  |  | 2.81 | <0.001 | 2.58 3.05 | NA |  |  | NA |  |  | NA |  |  | 2.36 | <0.001 | 2.16 2.57 |
| Anosmia | NA |  |  | 1.88 | 0.006 | 1.20 2.96 | NA |  |  | NA |  |  | NA |  |  | 1.97 | 0.003 | 1.25 3.09 |
| Change in skin sensation | NA |  |  | 1.01 | 0.924 | 0.84 1.20 | NA |  |  | NA |  |  | NA |  |  | 0.91 | 0.306 | 0.76 1.09 |
| Constipation | NA |  |  | 1.83 | <0.001 | 1.68 2.00 | NA |  |  | NA |  |  | NA |  |  | 1.53 | <0.001 | 1.40 1.67 |
| Dizziness | NA |  |  | 1.34 | <0.001 | 1.24 1.45 | NA |  |  | NA |  |  | NA |  |  | 1.22 | <0.001 | 1.13 1.32 |
| **Sleep and psychiatric presentations** |  |  |  |  |  |  |  |  |  |  |  |  |  |  |  |  |  |  |
| Restless legs syndrome | NA |  |  | NA |  |  | 4.28 | <0.001 | 3.69 4.96 | NA |  |  | NA |  |  | 3.50 | <0.001 | 3.02 4.05 |
| Sleep apnea | NA |  |  | NA |  |  | 1.17 | 0.054 | 1.00 1.37 | NA |  |  | NA |  |  | 0.96 | 0.642 | 0.82 1.13 |
| Parasomnia | NA |  |  | NA |  |  | 1.36 | 0.024 | 1.04 1.77 | NA |  |  | NA |  |  | 1.06 | 0.670 | 0.81 1.38 |
| Depression | NA |  |  | NA |  |  | 2.41 | <0.001 | 2.25 2.59 | NA |  |  | NA |  |  | 1.79 | <0.001 | 1.66 1.93 |
| Schizophrenia | NA |  |  | NA |  |  | 4.34 | <0.001 | 3.69 5.11 | NA |  |  | NA |  |  | 3.45 | <0.001 | 2.93 4.07 |
| Bipolar disorder | NA |  |  | NA |  |  | 1.94 | <0.001 | 1.44 2.61 | NA |  |  | NA |  |  | 1.53 | 0.005 | 1.14 2.06 |
| **Comorbidities** |  |  |  |  |  |  |  |  |  |  |  |  |  |  |  |  |  |  |
| Epilepsy | NA |  |  | NA |  |  | NA |  |  | 2.96 | <0.001 | 2.59 3.37 | NA |  |  | 1.63 | <0.001 | 1.42 1.86 |
| Migraine | NA |  |  | NA |  |  | NA |  |  | 1.01 | 0.926 | 0.83 1.24 | NA |  |  | 0.84 | 0.095 | 0.69 1.03 |
| Gastroesophageal reflux disease | NA |  |  | NA |  |  | NA |  |  | 1.15 | 0.002 | 1.05 1.26 | NA |  |  | 0.95 | 0.299 | 0.87 1.04 |
| Gastritis | NA |  |  | NA |  |  | NA |  |  | 1.33 | <0.001 | 1.23 1.45 | NA |  |  | 1.03 | 0.505 | 0.95 1.12 |
| Crohn disease | NA |  |  | NA |  |  | NA |  |  | 1.04 | 0.879 | 0.60 1.80 | NA |  |  | 0.82 | 0.476 | 0.48 1.41 |
| Seborrheic dermatitis | NA |  |  | NA |  |  | NA |  |  | 1.29 | 0.002 | 1.10 1.52 | NA |  |  | 1.08 | 0.382 | 0.91 1.26 |
| **Risk factors** |  |  |  |  |  |  |  |  |  |  |  |  |  |  |  |  |  |  |
| Type 1 diabetes | NA |  |  | NA |  |  | NA |  |  | NA |  |  | 1.25 | 0.001 | 1.10 1.43 | 1.09 | 0.171 | 0.96 1.24 |
| Type 2 diabetes | NA |  |  | NA |  |  | NA |  |  | NA |  |  | 1.19 | <0.001 | 1.11 1.29 | 1.10 | 0.016 | 1.02 1.19 |
| Hypertension | NA |  |  | NA |  |  | NA |  |  | NA |  |  | 1.57 | <0.001 | 1.42 1.75 | 1.35 | <0.001 | 1.21 1.50 |
| Hypercholesterolemia | NA |  |  | NA |  |  | NA |  |  | NA |  |  | 1.11 | 0.004 | 1.04 1.19 | 0.99 | 0.881 | 0.93 1.07 |
| Traumatic brain injury | NA |  |  | NA |  |  | NA |  |  | NA |  |  | 2.21 | <0.001 | 1.85 2.65 | 1.53 | <0.001 | 1.28 1.83 |
| Alcohol misuse | NA |  |  | NA |  |  | NA |  |  | NA |  |  | 1.55 | <0.001 | 1.30 1.85 | 1.07 | 0.483 | 0.89 1.27 |

When adjusting for motor, sensory, and autonomic presentations of PD, the risk of PD was 43% (HR=0.57 [0.53-0.61]) lower in the second period than in the first period (Table 2, Model 2). Tremor (HR=11.31 [10.36-12.35]), gait impairment (HR=2.81 [2.58-3.05]), anosmia (HR=1.88 [1.20-2.96]), constipation (HR=1.83 [1.68-2.00]), and dizziness (HR=1.34 [1.24-1.45]) were significantly positively associated with PD risk.

Adjustment for sleep and psychiatric presentations revealed a 33% reduced risk of PD (HR=0.67 [0.62-0.72] in the second period (Table 2, Model 3). The presence of restless legs syndrome (HR=4.28 [3.69-4.96]), parasomnia (HR=1.36 [1.04-1.77]), depression (HR=2.41 [2.25-2.59]), schizophrenia (HR=4.34 [3.69-5.11]), and bipolar disorder (HR=1.94 [1.44-2.61]) was positively associated with the risk of PD.

When we adjusted the model for comorbidities (Model 4) or for risk factors (Model 5), we observed a significantly reduced risk of PD in the second period by 22% (Model 4: HR=0.78 [0.73-0.84]) or 23% (Model 5: HR=0.77 [0.72-0.83]). The following comorbidities and risk factors were significantly positively associated with PD: epilepsy (HR=2.96 [2.59-3.37]), gastroesophageal reflux disease (HR=1.15 [1.05-1.26]), gastritis (HR=1.33 [1.23-1.45]), seborrheic dermatitis (HR=1.29 [1.10-1.52]), type 1 diabetes (HR=1.25 [1.10-1.43]), type 2 diabetes (HR=1.19 [1.11-1.29]), hypertension (HR=1.57 [1.42-1.57]), hypercholesterolemia (HR=1.11 [1.04-1.19]), traumatic brain injury (HR=2.21 [1.85-2.65]), and alcohol abuse (HR=1.55 [1.30-1.85]).

The full model showed a 49% (HR=0.51 [0.47-0.55]) reduction in the risk of PD in the second period as compared to the first period (Table 2, Model 6). After adjusting for all diseases and conditions, parasomnia, gastroesophageal reflux disease, gastritis, seborrheic dermatitis, type 1 diabetes, hypercholesterolemia, and alcohol misuse were no longer associated with the risk of PD.

The sensitivity analysis with the competing-risk model accounting for the competing event of death revealed a 48% (SHR=0.52 [0.48-0.56]) reduction in the risk of PD in the second period as compared to the first period (Table 3, Model 7), suggesting that potential changes in mortality within the 10 years do not affect the Cox model results.

**Table 3:** Results of competing-risk regression models for the risk of Parkinson’s disease (PD) in terms of sub-hazard ratios (SHR). Source AOK 2004-2009, AOK 2014-2019.

|  |  |  | Model 7 |  |
| --- | --- | --- | --- | --- |
| Variable | SHR | P value | 95% CI |  |
| Period (Ref.: 2006-2008) |  |  |  |  |
| 2016-2018 | 0.52 | 0.000 | 0.48 | 0.56 |
| Sex (Ref.: Women) |  |  |  |  |
| Men | 1.84 | 0.000 | 1.71 | 1.98 |
| Age | 0.99 | 0.024 | 0.98 | 1.00 |
| Age<sup>2</sup> | 0.997 | 0.000 | 0.996 | 0.997 |
| Motor, sensory, and autonomic presentations |  |  |  |  |
| Tremor | 9.90 | 0.000 | 8.98 | 10.92 |
| Gait impairment | 2.22 | 0.000 | 2.02 | 2.45 |
| Anosmia | 2.07 | 0.002 | 1.30 | 3.28 |
| Change in skin sensation | 0.94 | 0.481 | 0.78 | 1.12 |
| Constipation | 1.44 | 0.000 | 1.31 | 1.58 |
| Dizziness | 1.27 | 0.000 | 1.17 | 1.37 |
| Sleep and psychiatric presentations |  |  |  |  |
| Restless legs syndrome | 3.60 | 0.000 | 3.07 | 4.21 |
| Sleep apnea | 0.98 | 0.823 | 0.83 | 1.15 |
| Parasomnia | 1.04 | 0.767 | 0.79 | 1.39 |
| Depression | 1.78 | 0.000 | 1.65 | 1.93 |
| Schizophrenia | 3.39 | 0.000 | 2.84 | 4.05 |
| Bipolar disorder | 1.57 | 0.005 | 1.15 | 2.15 |
| Comorbidities |  |  |  |  |
| Epilepsy | 1.54 | 0.000 | 1.33 | 1.77 |
| Migraine | 0.86 | 0.153 | 0.70 | 1.06 |
| Gastroesophageal reflux disease | 0.97 | 0.460 | 0.88 | 1.06 |
| Gastritis | 1.01 | 0.747 | 0.93 | 1.10 |
| Crohn disease | 0.82 | 0.497 | 0.47 | 1.44 |
| Seborrheic dermatitis | 1.11 | 0.206 | 0.94 | 1.31 |
| Risk factors |  |  |  |  |
| Type 1 diabetes | 1.09 | 0.185 | 0.96 | 1.24 |
| Type 2 diabetes | 1.06 | 0.117 | 0.98 | 1.15 |
| Hypertension | 1.35 | 0.000 | 1.21 | 1.51 |
| Hypercholesterolemia | 1.02 | 0.635 | 0.95 | 1.09 |
| Traumatic brain injury | 1.47 | 0.000 | 1.22 | 1.77 |
| Alcohol misuse | 1.02 | 0.812 | 0.85 | 1.23 |

Ref.: Reference, SHR: Sub hazard ratio, CI: Confidence interval

## Discussion

In Western aging societies, epidemiologists have observed an increase in the incidence and prevalence of non-communicable diseases ^10,11^. Hence one could assume that the prevalence and incidence of PD should increase over time in Western aging societies, mainly with increasing life expectancy, now reaching 80 years in the EU in 2020 according to Eurostat (www.ec.europa.eu). In fact, PD incidence is increasing globally with an aging population and in males, with some regional variations ^12^. Accordingly, we also found an increase in PD incidence rates with increasing age in the German population studied. However, this finding was accompanied by an overall decrease in the age-specific incidence rates of PD over a ten-year period in both sexes, even when controlling for sex, PD prodromal and risk factors, comorbidities, and changes in death rates. While all neuropsychiatric symptoms and associated comorbidities were associated with an increased risk of PD, they did not change the significant decline in PD incidence. Thus, in our study changes in prodromal diagnoses and PD risk factors cannot explain the time trend in PD incidence rate.

Data from two other national-level studies in Germany confirmed the decrease in the annual incidence of idiopathic PD by 25-30% for ages 50 and above in both sexes within the same observation period from to 2013 to 2019 ^6,7^. The publication of such studies left the neurology community unprepared and rather surprised^8^, although studies from other parts of the world have partly reported similar trends. A study in Estonia reported that the overall PD incidence has remained comparatively stable over the last 20 years ^13^. Another study conducted in Rotterdam, showed an overall decreased incidence of PD between 1990 and 2011 ^14^. Similar results have been reported for South Korea demonstrating that between 2012 and 2015, the incidence of PD gradually decreased ^15^. In a large cohort study from Ontario, Canada, the incidence of mid/late-onset Parkinsonism decreased by 13% between 1996 and 2014 ^16^. Several concerns regarding the missing mechanistic explanation and the pure epidemiological nature of the data have been raised in follow-up debates ^8^.

The pathophysiological reason for the decreased incidence of PD observed in Germany is not clear, as most Westernized countries report an increase ^12,17^, and only a few others show a decrease in PD incidence ^14^,^15^.

### Variations in the prevalence and incidence of PD are influenced by several factors

For instance, there is suspicion of similar geographical patterns of specific pesticide use and PD incidence. This refers to toxins, including paraquat, which has been banned in many countries but is still in use in greater quantities in the U.S. ^18^,^19^, a country that reports continuously increased PD rates, at least in a regional fashion ^20^. Interestingly, certain countries, such as England, which have banned the use of paraquat, continue exporting it oversees (U.S., Brazil, South Africa, Taiwan)^21^. Interestingly, a nationwide study conducted in France aimed to examine the association of PD incidence with pesticide expenditures^22^. A 16% higher PD incidence was identified in cantons with the highest pesticide expenditure for vineyards without designation of origin, characterized by high fungicide use. The industrial solvent chlorinated halocarbon trichloroethylene (TCE) is another neurotoxic chemical linked to PD. Trichloroethylene can contaminate the air, water, and soil, and although its usage has been banned by the European Union and two U.S. states, it is still permitted for certain purposes ^23^ and is detected in drinking water ^24^. Several studies in rodents have shown that TCE induces dopaminergic neurodegeneration^25,26^. TCE is also able to induce alterations in the gut microbiome in a manner that reflects microbial signatures observed in idiopathic PD, confirming that exposure to this chemical may influence PD etiology via the gut-brain axis ^27^. An epidemiological study conducted on twins confirmed a significant association between TCE exposure and PD risk ^28,29^. However, according to the abovementioned study, the interval from the time of exposure to PD diagnosis varies from 10 to 40 years. This delay complicates the identification of a direct incidental link between TCE and PD although new evidence further corroborates that association ^29^. So far, the varied levels of pesticide use and the interplay with other risk factors do not allow a correlation between pesticide use and PD incidence ^30^

Another dynamic factor may be the continuous industrialization of countries such as China, where the prevalence has doubled from 1999 to 2016 ^17^. Asian countries are catching up as they had previously lower prevalence rates due to the lower prevalence of certain pathogenic risk factors and the absence of age-related changes in nigrostriatal dopaminergic neurons ^20,31^. Additionally, socioeconomic factors may interact with genetic factors because differences in LRK2 mutations may contribute to variations across ethnic groups, even in sporadic PD ^32,33^. This fact is further complicated by the occurrence of age-related conditions in subjects with migration backgrounds in Western countries.

Other modulating factors obscuring the clear report of PD incidences might include the use of artificial light ^34^, inconsistent diagnostic criteria, or even consanguineous marriages, which are more prevalent in Middle Eastern countries, possibly responsible for the higher prevalence of PD in these populations ^35^.

The association between smoking and PD incidence might represent a good example of how the pathophysiology and socioeconomic factors interact. Some studies have suggested that nicotine and tobacco may have neuroprotective effects and reduce the risk of Parkinson’s disease ^36,37^. Based on this information, one might expect an increase in PD in Germany, where smoking prevalence has decreased recently, especially in the female population ^38^. A similar complicated association is likely for caffeine consumption^39^. Even more trivial is the growing role of the microbiome in PD pathophysiology, a contributing factor that is considerably affected by the environment ^40^.

Another interesting factor was the association between residential greenness and PD. In Metro Vancouver, Canada, more than 600,000 residents aged between 45 and 84 years participated in a research study that revealed the protective effects of greenness on PD ^41^.

Similar results were obtained in a study sample from Korea that took place between 2007 and 2015 ^42^, as well as in China between 2015 and 2018 ^43^. These studies demonstrate how the presence of greenness in residential areas may have a protective role in neurodegenerative disorders, and the degree to which governments should consider such effects in their public health and more general strategies.

Significant contributions may arise from the correlation between migration and PD. Age-related pathological conditions among subjects with a migration background and comprising ethnic minorities pose an underappreciated but emerging challenge for Westernized societies.

Despite the advantage of using routinely collected data (large sample sizes, low panel attrition, no selection bias, and no recall bias due to self-reported diseases and conditions), a thorough investigation of our data led us to identify several constraints to be considered. Data are routinely collected for reimbursement purposes and are subject to legal changes. Therefore, changes in the coding of diagnoses do not necessarily reflect changes at the epidemiological level. In our case, we do not assume declining awareness of PD across the two observation periods. Rather, the opposite would be expected given the introduction of chronic disease coding incentives in 2013 ^44^, which could lead to higher rates in the second period. In addition, we observed the time of the first diagnosis rather than the time of onset. Delayed diagnosis may partly explain lower rates in the second period.

However, a sensitivity analysis revealed only a non-significant increase from 4.8 to 6.5 months over the 10 years (results not shown) when the time between the diagnosis of at least two cardinal symptoms of possible PD and the first PD diagnosis was considered. A bias in disease burden could also be introduced by changes in birth cohort sizes; therefore, we controlled all our models for the age structure of both cohorts.

In conclusion, our study showed a decrease in PD incidence rates in most age groups in Germany and a shift towards higher ages of first diagnosis. Single risk factors and comorbidities remained positively associated with PD incidence. PD occurrence varies substantially across European countries, with higher rates in northern and lower rates in southern countries ^45^. However, no clear associations were identified to explain the observed patterns. Further research is needed to fully understand the relationship between Parkinson’s disease, its risk factors, and comorbidities, and how these affect trends in the occurrence of the disease.

## Acknowledgement

We are grateful to the Scientific Research Institute of the AOK, WIdO, for providing the data.

## Data availability

The scientific research institute of the AOK (WIdO) has strict rules regarding data sharing because of the fact that health claims data are a sensitive data source and have ethical restrictions imposed due to concerns regarding privacy. Anonymized data are available to all interested researchers upon request. Interested individuals or an institution who wish to request access to the health claims data of the AOK may contact the WIdO (webpage: http://www.wido.de/, mail).

**Table S1:**
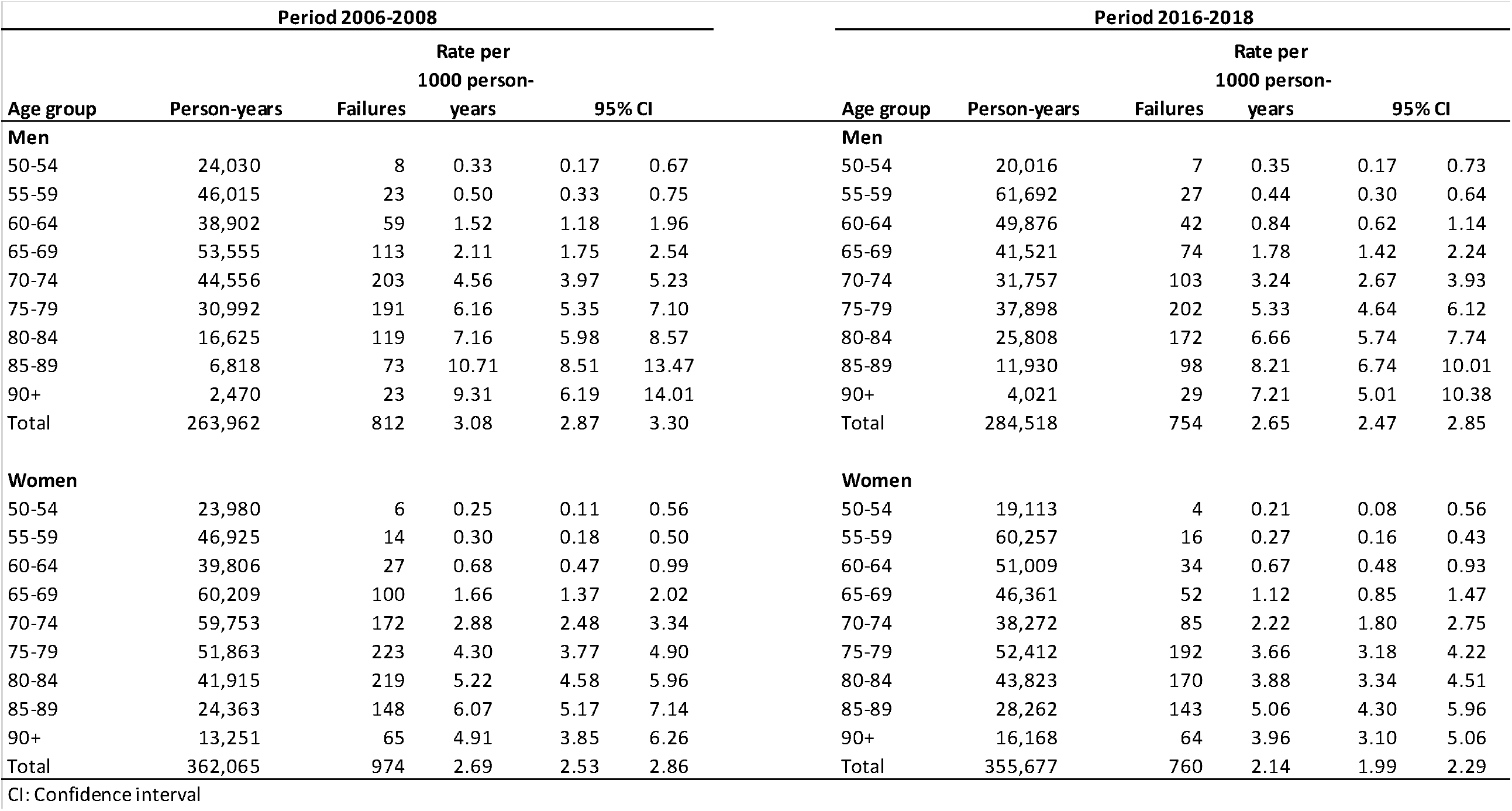
Results of incidence rates of Parkinson’s disease (PD) according to age groups. Source AOK 2004-2009, AOK 2014-2019.

